# Patient demographics, medical factors, treatment modalities and satisfaction at five traditional Chinese medicine practices: A cross-sectional study

**DOI:** 10.1101/2024.12.10.24318798

**Authors:** Yingchao Liu, Xiaoying Lyu, Saroj K. Pradhan, Yiming Li, Xingfang Liu, Ralf Bauder, Tanja Heggli, Xiaying Wang, Michael Furian

## Abstract

1

**BACKGROUND:** Traditional Chinese Medicine (TCM) is increasingly integrated into healthcare and insurance systems, therefore, it is essential to understand its current status and patients’ perspectives.

**METHODS:** This cross-sectional study was conducted from January 1st to December 31st, 2023, across five TCM practices in Switzerland. All patients attending their sixth therapy session were invited to complete an electronically anonymized questionnaire covering patient demographics, treatment experiences, and satisfaction.

**RESULTS:** A total of 461 patients participated in the survey, with the majority being female (60.1%) and aged 50 years or older (57.4%). Among them, 54.0% reported multiple health conditions, with 32.9% having musculoskeletal disorders and 31.8% suffering from chronic pain as the main reasons for seeking therapy. Most patients received weekly TCM treatments (91.3%), with 50.7% also undergoing conventional therapies. Of the respondents, 50.0% reported full coverage for their TCM therapy costs. TCM was mainly accessed through personal recommendations (44.5%), and 92.2% experienced wait times under 10 minutes. Acupuncture was the predominant treatment (95.7%), with 35.8% receiving additional dietary advice. Overall satisfaction reached 96.5%, and 99.5% expressed intent to continue TCM. Full compared to no TCM expense coverage was positively associated with treatment satisfaction (odds ratio = 2.42, 95% CI [1.10 to 5.31], p = 0.028), while other medical factors showed no significant impact on satisfaction.

**CONCLUSION:** This study indicates that women, patients over 50 years, and those with multiple health conditions, especially musculoskeletal and pain conditions, are more likely to seek regular integrated TCM treatment. Patients reported high satisfaction with TCM, and treatment satisfaction was positively associated with full compared to no health insurance coverage. Future research should include more TCM practices and patients to further generalize these findings and precisely assess the impact of insurance coverage on TCM satisfaction to meet patient needs better.

## 2 Background

Traditional Chinese Medicine (TCM) has a long history and includes therapy modalities such as acupuncture, cupping, herbal medicine, and life cultivation. In Europe, TCM is categorized as Complementary and Alternative Medicine (CAM). It has been recognized through including TCM herbal drugs in the *European Pharmacopoeia*[1] and achieved a milestone when TCM diagnostic patterns were included in the International Classification of Diseases (ICD) 11^th^ Revision, Chapter 26.[2, 3] A literature search in 2012 estimated at least 305,000 registered CAM providers in the European Union, acupuncture (n = 96,380) was the most widely therapeutic method for both medical (80,000) and non-medical (16,380) practitioners.[4] Research indicates that the usage of complementary medicine (CM) in Switzerland is comparable to that in other countries, such as Germany, the United Kingdom, the United States, and Australia. Thus, investigating the situation of TCM in Switzerland has significant relevance.[5]

Since 2012, in Switzerland, TCM has been covered by basic health insurance when provided by certified physicians; if provided by accredited therapists, it is covered by supplementary health insurance.[6] Consequently, the prevalence of CM use significantly increased between 2012 and 2017 from 24.7% to 28.9% (p<0.001), and compared to users of conventional healthcare, factors such as gender and age are independent determinants of CM use.[7] Cross-sectional surveys in representative samples of Swiss adult residents also showed a steady increase in the use of TCM from an initial 6.6% (use of acupuncture or TCM) in 2007 to 8.5% in 2017 (p <0.05 between 2007 vs. 2017).[5, 7]

In 2008, Michlig et al. conducted a prospective study in which patients reported significantly higher satisfaction after a 4-week follow-up when receiving TCM treatment from conventional physicians with additional TCM certification compared to conventional medicine therapy alone. However, there was no significant difference in complete recovery or marked symptom improvement.[8] Nevertheless, this study did not identify the TCM-related factors influencing treatment satisfaction. Since most TCM providers are not Western Medicine physicians, further research is warranted.

Although there are many studies on CAM from various databases, research on the real-world application of TCM and patient perspectives remains limited.[9, 10] Therefore, this cross-sectional study investigates patient characteristics, medical factors, treatment modalities, and satisfaction among individuals undergoing TCM treatment in five practices during the first year of implementing a satisfaction questionnaire.

## 3 Methods

### 3.1 Study design

This cross-sectional study was conducted between January 1 and December 31, 2023, across five TCM practices of the TCM Ming Dao Group AG in Switzerland (practices located in Bad Zurzach, Baden, Lenzburg, Wil, Zug). In agreement with the Swiss Human Research Act (HRA) and due to the assessment of anonymized data, this survey did not require ethical approval. This was confirmed by the local Ethics Committee Northwest and Central Switzerland (Req-2022-01098). The completion and findings of the individually completed questionnaires were not available to the TCM practice staff during the assessment period.

### 3.2 Study population and eligibility

The survey was distributed to German or English-speaking patients who signed the general consent[10] and attended the sixth therapy session, typically after three weeks of treatment, at one of the five TCM practices. This timing of the survey gave patients enough insight into the TCM treatment. Patients with less than six therapy sessions, who were unwilling to participate in the survey, or who had poor reading or language capability were excluded from the study.

### 3.3 Data collection and managem

The survey was created with the software EvaSys[11] and completed by the patients using a tablet during the waiting period for their subsequent treatment. An example of the survey is illustrated in the Supplement of this article. The survey encompassed a broad spectrum of data, including demographics (such as sex and age), medical factors (such as TCM practice, years of experience for TCM providers, treatment season, health conditions, treatment regularity, concurrent treatment with conventional medicine, and insurance status and coverage), TCM treatment modalities, and questions related to patients’ satisfaction.

The implemented questionnaire was created in collaboration with different TCM providers and senior researchers, and the feedback from the World Health Organization (WHO), Department of Complementary Medicine, Geneva, Switzerland.

### 3.4 Statistical analysis

The reporting of the findings of this study follows the Strengthening the Reporting of Observational Studies in Epidemiology (STROBE) guidelines (Supplement).[12] The data are summarized as numbers and proportions. Comparisons between proportions were conducted with χ^2^ tests. Ordered logistic regression analysis examined the relationship between treatment satisfaction, patient demographics, and medical factors. A two-sided p<0.05 was considered to reflect statistical significance. All statistical analyses were conducted with the software R, version 4.0.5 (R Foundation for Statistical Computing in Vienna, Austria).

## 4 Results

### 4.1 Study participation

Out of 1,095 patients starting a new TCM therapy in 2023, 611 (55.8%) signed the general consent form when starting the TCM therapy. Of these, 473 (43.2%) received a sixth TCM therapy session, and 461 anonymized satisfaction surveys were returned (Figure 1).

**Figure 1:**
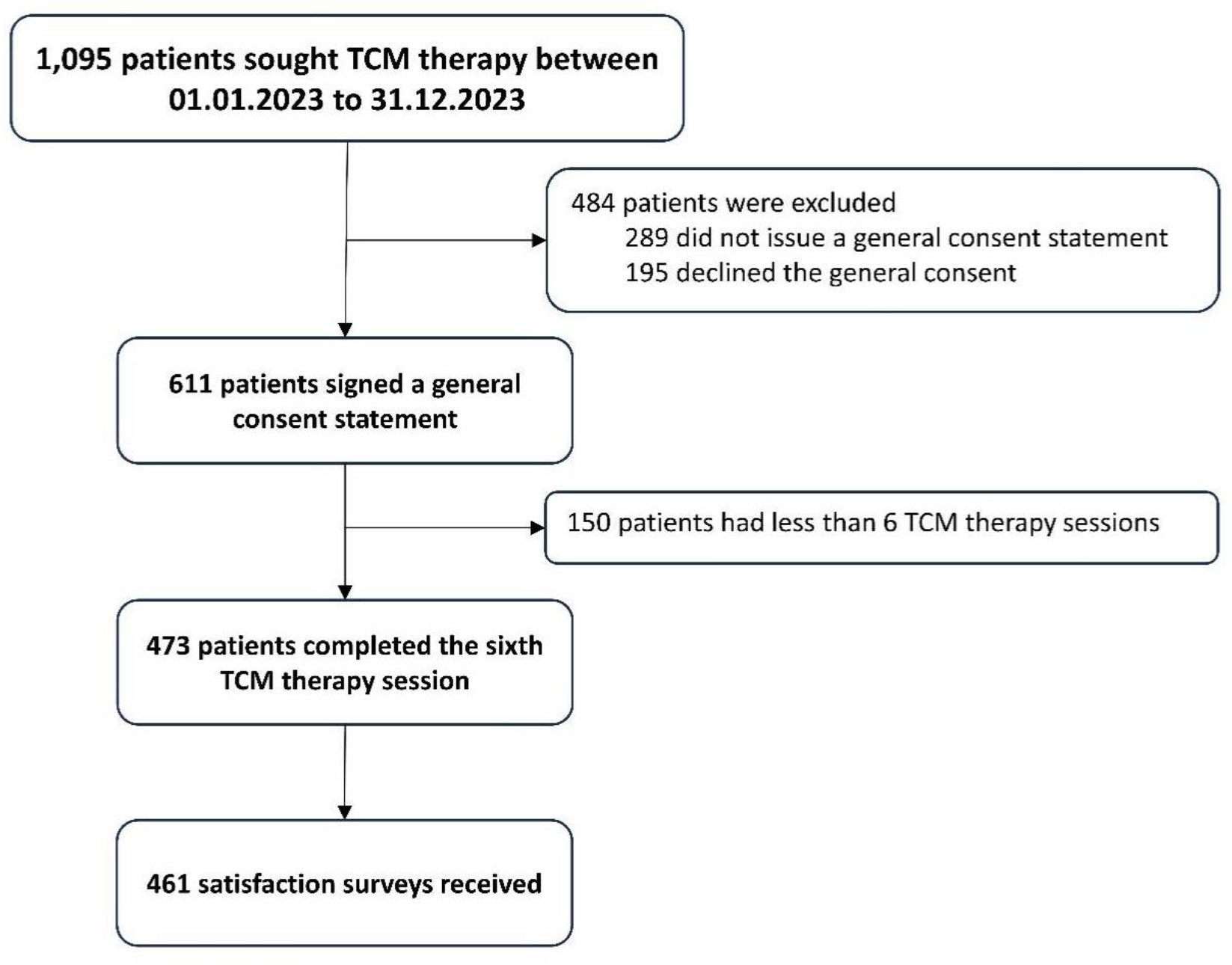
Study flow chart. Patients who consented to the general consent form were asked to participate in the satisfaction survey at their sixth traditional Chinese medicine therapy session. TCM, traditional Chinese medicine.

### 4.2 Patient and TCM practice characteristics

In the study, 277 (60.1%) were women, significantly more than men (p <0.001). Patients aged ≥50 years outnumbered those aged <50 years (p = 0.002). The median age category was 50 to 60 years, and the average age of male patients was higher than that of female patients (Table 1, Figure 2). In terms of treatment regularity, patients receiving weekly treatments (91.3%) significantly outnumbered those receiving treatments irregularly (p <0.001). Half of all respondents reported concurrent treatment with conventional medicine. There was a significant difference in insurance coverage for TCM treatment costs (p <0.001); 50% reported that they could cover the TCM treatment expenses fully, 45% partially, and 5% not at all.

**Table 1.**
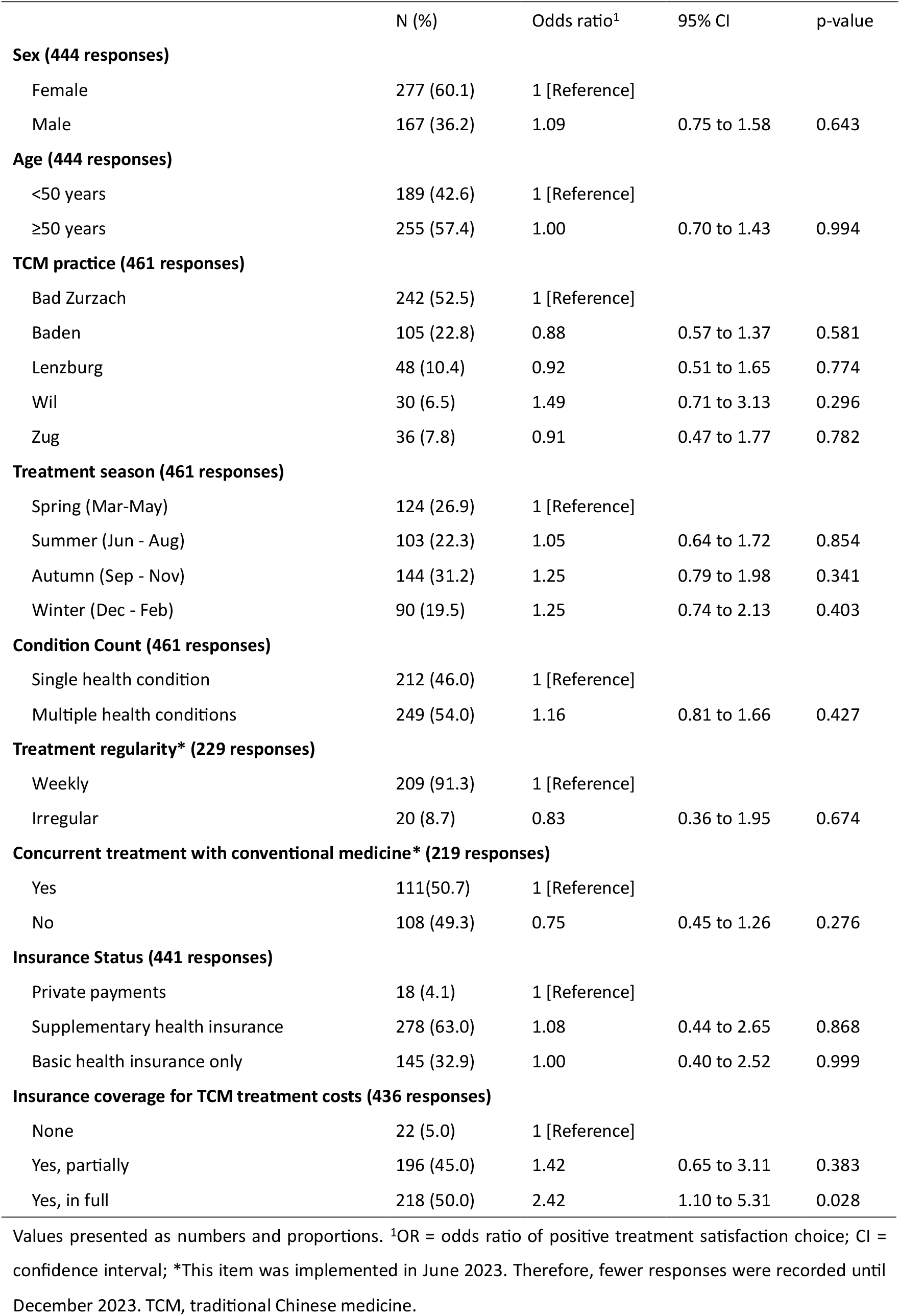
Characteristics of TCM practices and patients seeking TCM therapy.

|  | N (%) | Odds ratio<sup>1</sup> | 95% CI | p-value |
| --- | --- | --- | --- | --- |
| **Sex (444 responses)** |  |  |  |  |
| Female | 277 (60.1) | 1 [Reference] |  |  |
| Male | 167 (36.2) | 1.09 | 0.75 to 1.58 | 0.643 |
| **Age (444 responses)** |  |  |  |  |
| <50 years | 189 (42.6) | 1 [Reference] |  |  |
| ≥50 years | 255 (57.4) | 1.00 | 0.70 to 1.43 | 0.994 |
| **TCM practice (461 responses)** |  |  |  |  |
| Bad Zurzach | 242 (52.5) | 1 [Reference] |  |  |
| Baden | 105 (22.8) | 0.88 | 0.57 to 1.37 | 0.581 |
| Lenzburg | 48 (10.4) | 0.92 | 0.51 to 1.65 | 0.774 |
| Wil | 30 (6.5) | 1.49 | 0.71 to 3.13 | 0.296 |
| Zug | 36 (7.8) | 0.91 | 0.47 to 1.77 | 0.782 |
| **Treatment season (461 responses)** |  |  |  |  |
| Spring (Mar-May) | 124 (26.9) | 1 [Reference] |  |  |
| Summer (Jun - Aug) | 103 (22.3) | 1.05 | 0.64 to 1.72 | 0.854 |
| Autumn (Sep - Nov) | 144 (31.2) | 1.25 | 0.79 to 1.98 | 0.341 |
| Winter (Dec - Feb) | 90 (19.5) | 1.25 | 0.74 to 2.13 | 0.403 |
| **Condition Count (461 responses)** |  |  |  |  |
| Single health condition | 212 (46.0) | 1 [Reference] |  |  |
| Multiple health conditions | 249 (54.0) | 1.16 | 0.81 to 1.66 | 0.427 |
| **Treatment regularity\* (229 responses)** |  |  |  |  |
| Weekly | 209 (91.3) | 1 [Reference] |  |  |
| Irregular | 20 (8.7) | 0.83 | 0.36 to 1.95 | 0.674 |
| **Concurrent treatment with conventional medicine\* (219 responses)** |  |  |  |  |
| Yes | 111(50.7) | 1 [Reference] |  |  |
| No | 108 (49.3) | 0.75 | 0.45 to 1.26 | 0.276 |
| **Insurance Status (441 responses)** |  |  |  |  |
| Private payments | 18 (4.1) | 1 [Reference] |  |  |
| Supplementary health insurance | 278 (63.0) | 1.08 | 0.44 to 2.65 | 0.868 |
| Basic health insurance only | 145 (32.9) | 1.00 | 0.40 to 2.52 | 0.999 |
| **Insurance coverage for TCM treatment costs (436 responses)** |  |  |  |  |
| None | 22 (5.0) | 1 [Reference] |  |  |
| Yes, partially | 196 (45.0) | 1.42 | 0.65 to 3.11 | 0.383 |
| Yes, in full | 218 (50.0) | 2.42 | 1.10 to 5.31 | 0.028 |

Values presented as numbers and proportions. <sup>1</sup>OR = odds ratio of positive treatment satisfaction choice; CI = confidence interval; \*This item was implemented in June 2023. Therefore, fewer responses were recorded until December 2023. TCM, traditional Chinese medicine.

**Figure 2:**
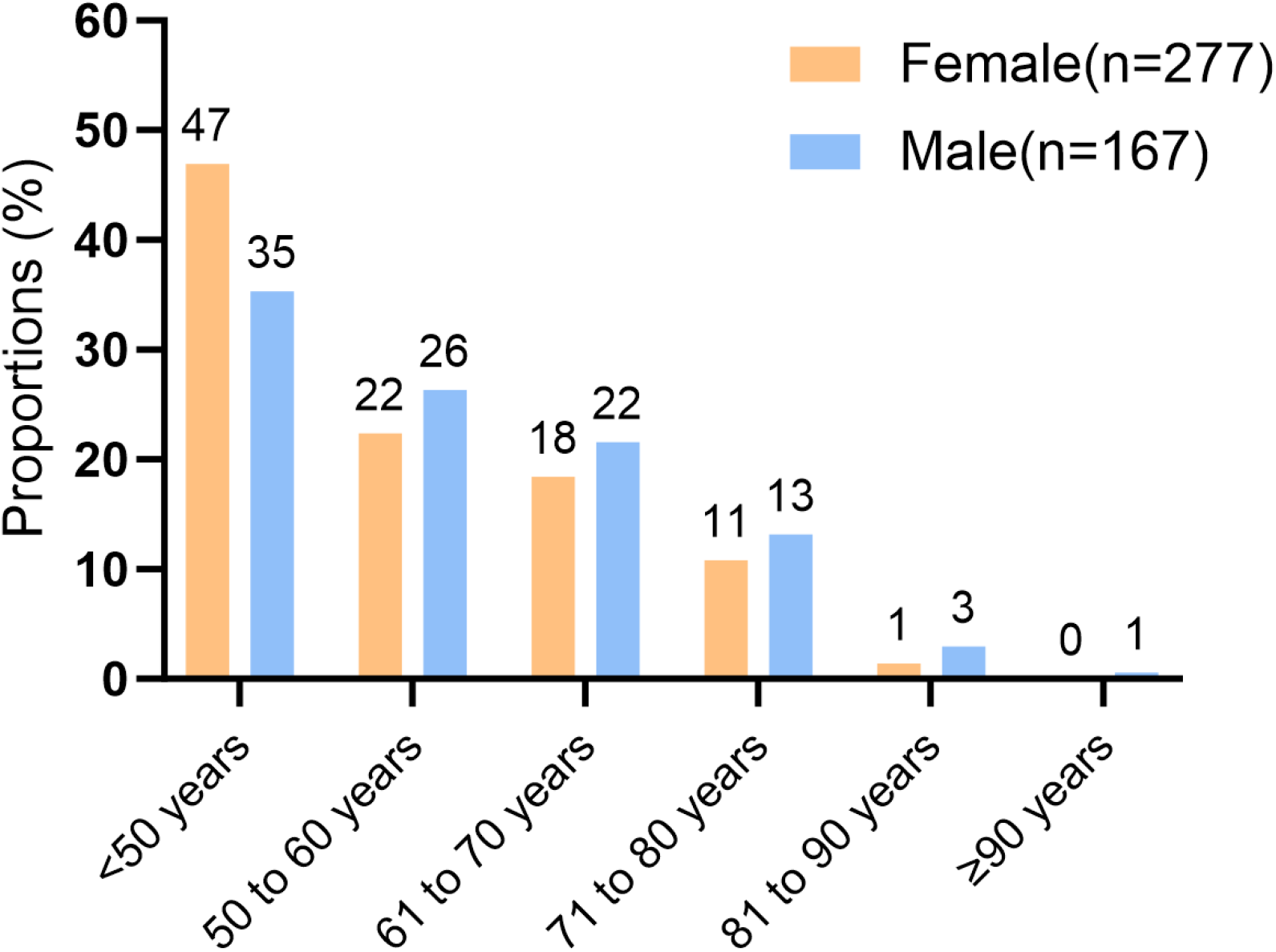
Age distribution in female and male returning a satisfaction questionnaire.

There were significant differences in the total number of enrolled patients among the five TCM practices (p <0.001), as well as among the four seasons, with winter having the fewest patients (19.5%) (Table 1). Regarding TCM providers, four held bachelor’s degrees, two held master’s degrees, and four held doctoral degrees in TCM. Their average years of TCM practice was 22.7 ± 11.4 years, ranging from 10 to 40 years.

### 4.3 Medical factors

The survey listed 23 possible health conditions for seeking TCM treatment. Each patient reported an average of 2.0 ± 1.5 different health conditions, ranging from 1 to 9. Patients with multiple conditions (54.0%) outnumbered those with a single condition (46.0%), although the difference was not statistically significant. Among all patients, the most reported reasons for seeking TCM treatment were musculoskeletal disorders (33%) and chronic pain conditions (32%) (Figure 3, Panel A). Both acute and chronic pain conditions were closely linked with musculoskeletal disorders (Figure 3, Panels B to D).

**Figure 3:**
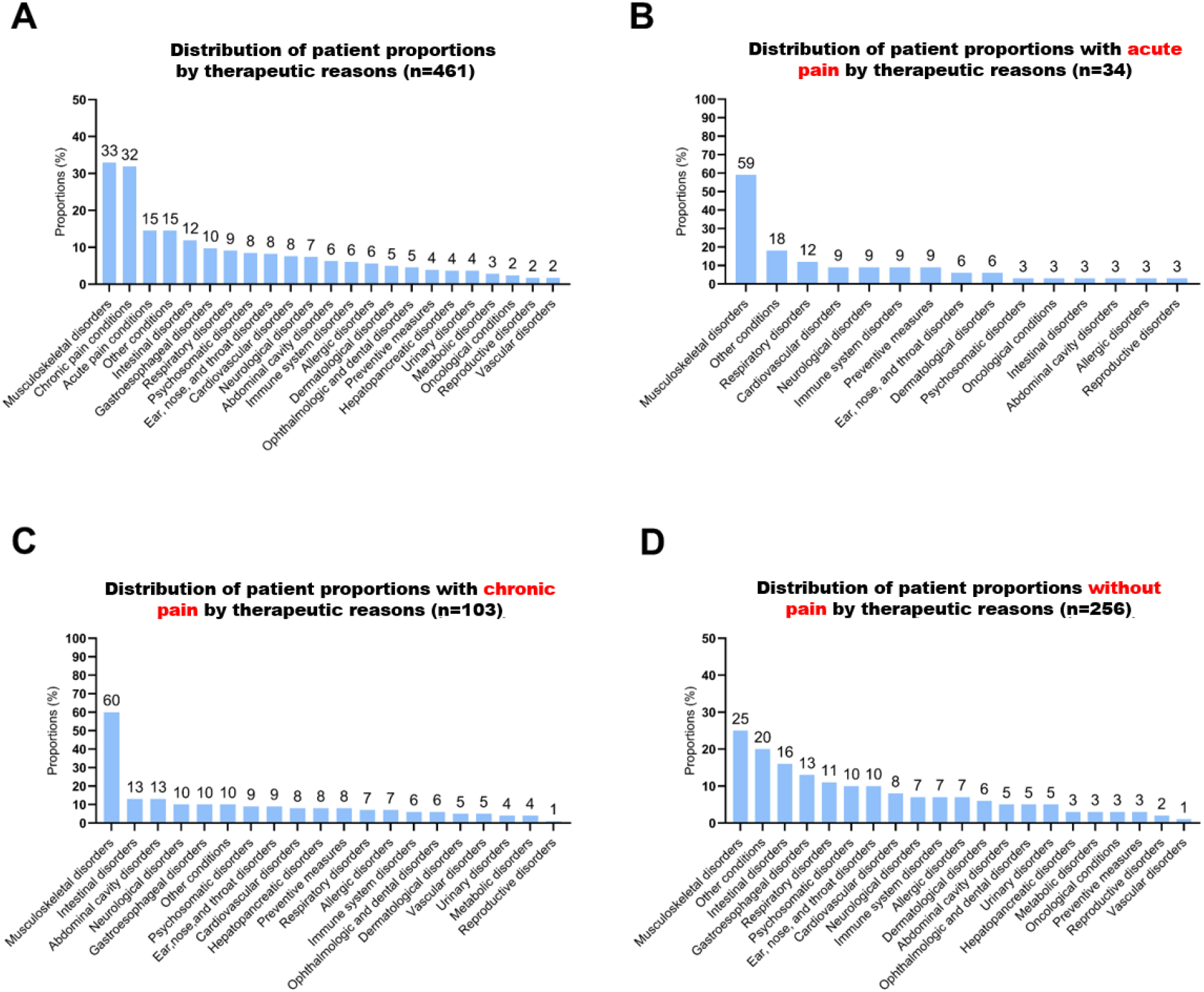
Distribution of patient proportions by therapeutic reasons. Panel A shows the proportion of overall therapeutic reasons; Panel B shows the proportion of therapeutic reasons under acute pain; Panel C shows the proportion of therapeutic reasons under chronic pain; Panel D shows the proportion of therapeutic reasons without pain.

### 4.4 TCM treatment modalities

Patients were primarily introduced to TCM through personal recommendations (45%) and the Internet (23%). Most experienced minimal waiting times before therapy, with over 50% reporting no wait. The patients reported an average of 1.5 ± 0.4 different treatment therapies and 1.3 ± 0.1 different lifestyle modification recommendations. Acupuncture was the most used therapy in more than 95% of cases. Many patients received lifestyle advice, particularly on diet. Overall, 99.5% of patients were positive about continuing TCM treatment (Table 2).

**Table 2.** TCM treatment modalities.

| **Parameters** | **n (%)** |
| --- | --- |
| **How did you first learn about TCM? (461 responses)** |  |
| Through recommendation | 205 (44.5) |
| Through the internet | 104 (22.6) |
| Through prior treatment | 85 (18.4) |
| Through medical referral | 74 (16.0) |
| Through an event | 30 (6.5) |
| Through advertisement | 16 (3.5) |
| Through other reasons | 13 (2.8) |
| Through social media | 2 (0.4) |
| **Waiting time before therapy (448 responses)** |  |
| No Waiting Time | 243 (54.2) |
| Up to 5 Min | 128 (28.6) |
| Up to 10 Min | 42 (9.4) |
| Up to 20 Min | 18 (4.0) |
| Up to 30 Min | 13 (2.9) |
| More than 30 Min | 4 (0.9) |
| **Applied TCM treatment therapy (461 responses)** |  |
| Acupuncture | 441 (95.7) |
| Cupping | 135 (29.3) |
| Acupressure and massage | 65 (14.1) |
| Chinese herbal medicine | 19 (4.1) |
| Moxibustion | 5 (1.1) |
| Other Therapy | 4 (0.9) |
| **Additional advice on lifestyle modification (461 responses)** |  |
| Diet | 165 (35.8) |
| Other recommendations | 123 (26.7) |
| Sleep hygiene | 94 (20.4) |
| Physical activity | 91 (19.7) |
| No specific recommendations | 90 (19.5) |
| Clothing and thermal regulation | 60 (13.0) |
| **Continue of TCM treatment\* (227 responses)** |  |
| Definitely yes | 166 (73.1) |
| Rather yes | 60 (26.4) |
| Rather no | 1 (0.4) |
| Definitely no | 0 (0.0) |

n = number of responses; \* This item was implemented in June 2023. Therefore, fewer responses were recorded until December 2023. TCM, traditional Chinese medicine.

### 4.5 Perceived satisfaction

Overall satisfaction of all respondents was 96.5% and similar across multiple domains, including treatment satisfaction, medical environment, administration, and TCM providers (Table 3, Figure 4). Treatment satisfaction tied to the perceived effectiveness of therapy was 86.3%, with 52.3% of respondents reporting they were “very satisfied” and 34.0% “satisfied”, while 5.7% were dissatisfied, and 8.0% were unable to evaluate. Other domains, such as billing inquiry, medical consultation, disease explanation, and treatment instruction, received “very satisfied” ratings of 65.9%, 69.4%, 60.4%, and 60.7%, respectively (Table 3, Figure 4).

**Table 3.** Satisfaction with various aspects of the TCM practices and treatment.

|  | Very Satisfied | Satisfied | Dissatisfied | Very Dissatisfied | Unable to evaluate |
| --- | --- | --- | --- | --- | --- |
| **Overall satisfaction** | NA | 96.5% | 0.9% | NA | 2.6% |
| **Treatment satisfaction** | 52.3% | 34.0% | 5.0% | 0.7% | 8.0% |
| **Medical environment** |  |  |  |  |  |
| Hygiene and neatness | 93.8% | 5.4% | 0.2% | 0.7% | 0.0% |
| Clinic atmosphere | 79.1% | 19.4% | 0.9% | 0.5% | 0.2% |
| Peace and relaxation | 81.5% | 15.8% | 2.0% | 0.7% | 0.0% |
| Patient privacy | 75.1% | 21.9% | 2.1% | 0.7% | 0.2% |
| **Administration** |  |  |  |  |  |
| Registration process | 90.9% | 8.0% | 0.0% | 1.1% | 0.0% |
| Reception courtesy | 93.9% | 5.0% | 0.0% | 0.9% | 0.2% |
| Billing inquiry | 65.9% | 24.6% | 1.4% | 0.5% | 7.7% |
| Clinic accessibility | 78.7% | 14.5% | 1.2% | 0.7% | 4.9% |
| **TCM provider** |  |  |  |  |  |
| Resonance perception | 87.5% | 11.4% | 0.2% | 0.9% | 0.0% |
| Care during treatment | 86.1% | 11.3% | 0.4% | 0.9% | 1.3% |
| Dedicated time | 82.7% | 14.9% | 1.8% | 0.7% | 0.0% |
| Medical consultation | 69.4% | 26.0% | 3.0% | 0.5% | 1.1% |
| Disease explanation | 60.4% | 31.9% | 3.7% | 0.5% | 3.5% |
| Treatment instruction | 60.7% | 30.2% | 3.4% | 0.7% | 5.0% |

Values represent proportions. TCM, traditional Chinese medicine; NA, not assessed.

**Figure 4:**
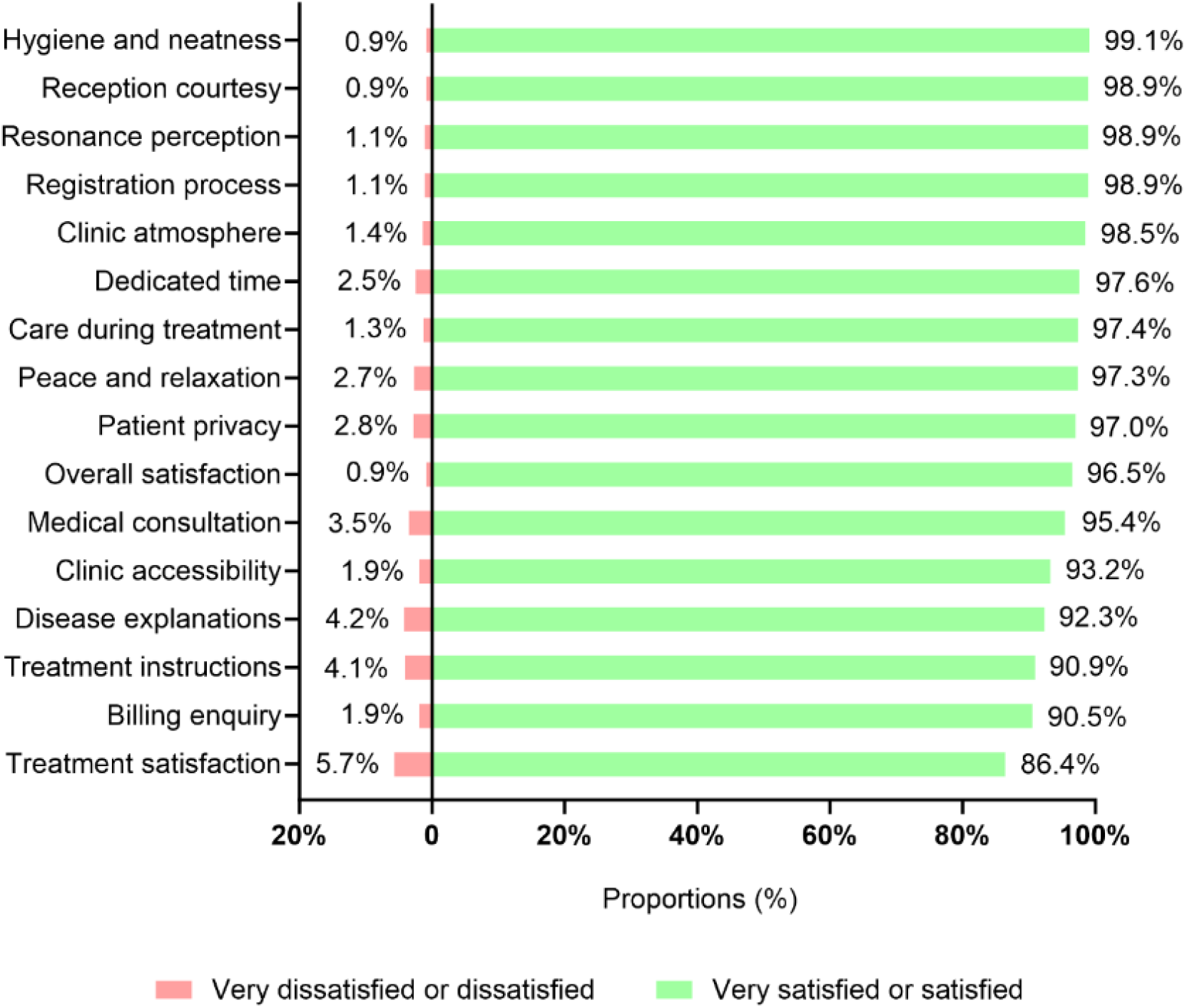
Patients’ satisfaction with various TCM practices and treatment aspects. TCM, traditional Chinese medicine.

In univariate ordered logistic regression analyses, full insurance coverage for TCM treatment costs was positively associated with higher treatment satisfaction (Table 1). Other variables did not show relevant associations with treatment satisfaction (Table 1).

## 5 Discussion

This cross-sectional study conducted in five Swiss TCM practices in the year 2023 revealed that patients seeking therapy suffer from a broad spectrum of underlying health conditions but report high satisfaction and perceived treatment effects after the sixth TCM therapy session. This description of patient demographics provides insights into the spectrum of TCM-treated health conditions and paves the way for more sophisticated, practice-based prospective studies, investigating TCM therapies and their effects on patients’ perceived and objective treatment effects.

The higher proportion of women seeking TCM therapy aligns with broader patterns observed in healthcare utilization worldwide, where women tend to use CM, including TCM, more frequently than men.[5, 13] Previous studies have indicated that this trend may be related to a higher prevalence of chronic diseases among women.[14] Similarly, the utilization of TCM therapy by different age groups reflects older adults, who are more likely to have pain or experience age-related declines in health, tend to use TCM more frequently than younger individuals (Figure 2).[15] TCM addresses both physical and emotional aspects of health, providing comprehensive care that targets medical conditions and supports emotional well-being, which may complement conventional healthcare practices.[16]

Patients seeking TCM treatment often present themselves with complex medical histories, posing both challenges and opportunities for TCM. This study’s most commonly treated conditions included musculoskeletal disorders, pain, and intestinal and gastroesophageal disorders. This aligns with previously established areas of strength for TCM based on evidence-based studies.[17] The American College of Physicians’ clinical practice guideline on noninvasive treatments for acute, subacute, and chronic low back pain recommends prioritizing non-drug approaches, such as exercise, multidisciplinary rehabilitation, and acupuncture.[18] Additionally, acupuncture has shown positive clinical trial results in treating gastroesophageal reflux disease, ulcerative colitis, and Crohn’s disease.[19-21] More prospective clinical research and multidisciplinary collaboration are needed to better define TCM’s role and value in contemporary healthcare.[22]

The previously mentioned demographic and medical factors, including TCM practices, treatment seasons, and the years of practice of TCM providers, showed no correlation with treatment satisfaction, which should be confirmed by other cross-sectional studies. In contrast, insurance coverage, full vs. no coverage for TCM treatment costs, was positively associated with higher treatment satisfaction. This is in accordance with other studies showing that economic factors may play a significant role in treatment satisfaction.[23] This financial burden can discourage patients from continuing treatment, even if they perceive positive health effects. Future research could inform policy adjustments to improve TCM accessibility and reduce financial strain on patients.

In this study, acupuncture was the most frequently used TCM treatment modality, with 95.7% of patients receiving it, which aligns with both existing literature and clinical practice norms.[24] According to the 2017 Swiss Health Survey of 18,832 patients, acupuncture was the most widely used TCM therapy within the past year.[25] Acupuncture is the most commonly chosen TCM treatment for musculoskeletal disorders and pain due to its proven efficacy.[17] Additionally, TCM emphasizes combining various treatment modalities to restore and balance the body. Patient satisfaction with TCM treatments is a complex and multidimensional outcome influenced by various factors.[26] The study found that while patients value the medical environment and administration significantly, treatment satisfaction has not been fully met. Enhancing rapid information about the billing and therapy cost coverage may improve adherence among patients with insufficient insurance coverage, indirectly boosting treatment satisfaction. Additionally, dedicating sufficient time to improve medical consultation, disease explanation, and treatment instruction may enhance treatment efficacy. Furthermore, treatment satisfaction varies across different disease types. Whether these differences in satisfaction are related to the timing of the survey, the inefficiency of the TCM therapy, or other factors could be addressed by prospective clinical trials within the TCM practices.

This study has several limitations. Firstly, the research relies on self-reported questionnaires, which may introduce reporting bias, especially as patients might give higher satisfaction ratings due to social expectations. Secondly, surveying the sixth TCM treatment session may lead to selection bias, as more satisfied patients are more likely to continue treatment. However, conducting the survey earlier than the sixth session would not have given the patients enough time to rate their satisfaction in various domains. Additionally, patients’ health conditions were not classified according to the International Classification of Diseases (ICD-10), which limits the accurate assessment of health status and may overlook important factors affecting satisfaction. Furthermore, since the study was conducted in Switzerland, the external validity of the results should be confirmed by other studies in other TCM practices. Future research should be conducted in a broader cultural and economic context to validate the applicability of the findings from this study.

## 6 Conclusion

This cross-sectional survey in five TCM practices at the sixth TCM therapy session revealed that most patients reported a high satisfaction related to the perceived treatment effect. The majority of patients were women and reported a broad range of health conditions, with musculoskeletal complaints as the most often reported health issue. Most often, the TCM treatment included acupuncture combined with lifestyle recommendations. Notably, patients reported high satisfaction in various TCM practices-related domains, i.e., hygiene, registration procedure or clinic atmosphere. Future research should prioritize practice-based, real-world studies that assess the effects of TCM treatments as well as the frequency of therapy abortion in relation to patients’ initial health conditions and disease burden. This approach will provide clearer evidence of TCM’s effectiveness and help guide its application in healthcare.

## Supporting information

STROBE_Checklist

Satisfaction_Questionnaire_ENG

## Data Availability

Data underlying this study can be requested by qualified researchers providing an approved proposal.

## 8 Acknowledgements

We thank the TCM Ming Dao and all patients for supporting this project.

## 8.1 Authors’ contributions

Conceptualization: Saroj K.Pradhan, Yiming Li, Xingfang Liu, Ralf Bauder and Michael Furian; data collection: Saroj K.Pradhan, Yiming Li, Ralf Bauder and Michael Furian; data analysis: Yingchao Liu, Xiaoying Lyu, Tanja Heggli, Xiaying Wang and Michael Furian; writing original draft: Yingchao Liu, Xiaoying Lyu and Michael Furian; Critical review of manuscript: all authors.

### 8.2 Financial Disclosure

This project received no specific funding from public, commercial, or non-profit funding agencies.

