## Supplementary material for "Patient demographics, medical factors, treatment modalities and satisfaction at five traditional Chinese medicine practices: A cross-sectional study": Satisfaction_Questionnaire_ENG

Dear patient Your health is important to us. To ensure that you feel comfortable, it is our highest desire to continuously improve and monitor our services. For this reason, we would be pleased when you spend 10 minutes and fill out our questionnaire. All obtained information will be anonymized, handled with uttermost discretion and will be only accessible to authorized internal personnel of the TCM Ming Dao AG. Your answers will not have any consequences on the physician-client relationship and your current and future TCM treatments. If you cannot or do not want to answer a question, you can move on to the next question. In case of minors or persons under the age of majority, this applies mutatis mutandis to their legal representatives.

### 1. Your team from TCM Ming Dao

1.1 At which location are you being treated?

- ○ Bad Zurzach
- ○ Baden
- ○ Basel
- ○ Lenzburg
- ○ Wil
- ○ Zug

### 2. Questions about your person

2.1 Please state your gender

- ○ Female
- ○ Male

2.2 Please tell us your age (female):

- ○ under 7 years
- ○ between 8 and 14 years
- ○ between 15 and 21 years
- ○ between 22 and 35 years
- ○ between 36 and 49 years
- ○ between 50 and 60 years
- ○ between 61 and 70 years
- ○ between 71 and 80 years
- ○ between 81 and 90 years
- ○ older than 90 years

2.3 Please tell us your age (male):

- ○ under 8 years
- ○ between 9 and 16 years
- ○ between 17 and 24 years
- ○ between 25 and 40 years
- ○ between 41 and 49 years
- ○ between 50 and 60 years
- ○ between 61 and 70 years
- ○ between 71 and 80 years
- ○ between 81 and 90 years
- ○ older than 90 years

2.4 How did you find out about us? Multiple answers possible.

- ☐ Events (e.g. trade fair, information evening, open day)
- ☐ Advertising (e.g. newspaper)
- ☐ Referral (e.g. family doctor, specialist, clinic)
- ☐ Recommendation by friends
- ☐ I have been treated here before
- ☐ Internet (e.g. Google, website TCM Ming Dao)
- ☐ Social media (e.g. YouTube, Facebook, Instagram)
- ☐ Other reasons

2.5 If you have chosen "other reasons", please tell us how you found us.

(maximum 200 characters)

2.6 Have you already completed this satisfaction questionnaire in the past?

- ○ Yes
- ○ No

2.7 If you have already completed a satisfaction questionnaire. How long ago was that?

- ○ Approx. 3 months
- ○ Approx. 6 months
- ○ Approx. 1 year
- ○ Longer than 1 year
- ○ Don't know anymore

2.8 Please tell us your complaints or the reason for the TCM treatment (optional). Multiple answers possible.

- [ ] Eyes, mouth
- [ ] Throat, nose, ears
- [ ] Heart, circulation
- [ ] Respiratory tract, lungs
- [ ] Stomach, oesophagus
- [ ] Pancreas, liver
- [ ] Lower abdomen
- [ ] Bowel
- [ ] Bladder, kidneys
- [ ] Joints, spine
- [ ] Arteries, veins
- [ ] Skin
- [ ] Pain (acute, up to 3 months)
- [ ] Pain (chronic, longer than 3 months)
- [ ] Metabolism (blood sugar, cholesterol)
- [ ] Infertility
- [ ] Psychosomatic disorder
- [ ] Immune system
- [ ] Allergy
- [ ] Neurological disease
- [ ] Cancer
- [ ] Life cultivation
- [ ] Other reasons

2.9 If you have chosen "other reasons", please tell us your complaints or the reason for the TCM treatment.

(maximum 200 characters)

2.10 How often do you receive TCM therapy?

○ Regularly (weekly) ○ Irregular ○ Seldom

2.11 Are you going to continue with TCM treatment?

○ Certainly yes ○ Rather yes ○ Rather not ○ Certainly not

2.12 Are you also being treated with conventional medicine?

○ Yes ○ No

2.13 How are you covered by health insurance?

○ Basic insurance ○ Supplementary insurance

○ Private (cash payer)

2.14 Can you bill your health insurance for the TCM treatment(s)?

○ Yes, fully ○ Yes, partly ○ No

2.15 How important is the qualification of the TCM therapist to you?

○ Very important ○ Important ○ Not that important

○ Not important ○ I don't know

### 3. Questions about TCM Secretariat and/or TCM Assistance

How is your overall satisfaction regarding...

| | very satisfied | rather satisfied | rather dissatisfied | very dissatisfied | not specified |
|---|---|---|---|---|---|
| 3.1 ... a smoothly check-in / registration process run? | ○ | ○ | ○ | ○ | ○ |
| 3.2 ... beeing treated by clerks and receptionist with courtesy and respect? | ○ | ○ | ○ | ○ | ○ |
| 3.3 ... advice on the possibilities of billing your insurance company? | ○ | ○ | ○ | ○ | ○ |
| 3.4 ... locating the TCM clinic (were the information provided by the clerks, receptionist, website helpful)? | ○ | ○ | ○ | ○ | ○ |

3.5 Average waiting time is about:

- ○ no waiting time
- ○ up to 5 min.
- ○ up to 10 min.
- ○ up to 20 min.
- ○ up to 30 min.
- ○ more than 30 min.

### 4. Consultation, explanation, treatment and attention from your TCM physician

4.1 I have so far received the following information about improving lifestyle habits. Multiple answers possible.

- ☐ Nutrition
- ☐ Sleep
- ☐ Clothing
- ☐ Exercise (e.g. Qi Gong / Tai Chi)
- ☐ No informations
- ☐ Other informations

4.2 If you have chosen "Other information", please tell us about the ways to improve your lifestyle habits.

(maximum 200 characters)

4.3 I have been treated with one or more of the following therapies:

- ☐ Acupuncture
- ☐ Chinese Herbal Therapy
- ☐ Cupping
- ☐ Moxa-Therapy
- ☐ Acupressure (Tuina massage)
- ☐ Other therapy
- ☐ I have not received any therapy yet

4.4 If you have selected "Other therapy", please name the therapy you were treated with.

(maximum 200 characters)

How is your overall satisfaction regarding...

| | very satisfied | rather satisfied | rather dissatisfied | very dissatisfied | not specified |
|---|---|---|---|---|---|
| 4.5 ... of his empathy? | ○ | ○ | ○ | ○ | ○ |
| 4.6 ... the time he spent with you? | ○ | ○ | ○ | ○ | ○ |
| 4.7 ... the general (medical) advice related to your situation? | ○ | ○ | ○ | ○ | ○ |
| 4.8 ... the information about the character, extent and severity of your complaints/illness? | ○ | ○ | ○ | ○ | ○ |
| 4.9 ... the possible therapeutic actions and their respective benefits and risks in the context of your treatment? | ○ | ○ | ○ | ○ | ○ |

### 5. Cleanliness, hygiene and privacy

What is your overall satisfaction in the TCM clinic regarding...

| | very satisfied | rather satisfied | rather dissatisfied | very dissatisfied | not specified |
|---|---|---|---|---|---|
| 5.1 ... hygiene, cleanliness and organisation? | ○ | ○ | ○ | ○ | ○ |
| 5.2 ... the atmosphere (e.g. furnishings, lighting, indoor air)? | ○ | ○ | ○ | ○ | ○ |
| 5.3 ... silence and relaxation? | ○ | ○ | ○ | ○ | ○ |
| 5.4 ... privacy? | ○ | ○ | ○ | ○ | ○ |

### 6. Overall satisfaction

How is your overall satisfaction regarding...

| | very satisfied | rather satisfied | rather dissatisfied | very dissatisfied | not specified |
|---|---|---|---|---|---|
| 6.1 ... the care during the treatment? | ○ | ○ | ○ | ○ | ○ |
| 6.2 ... the success of the treatment? | ○ | ○ | ○ | ○ | ○ |

6.3 Would you recommend the TCM clinic to others? ○ Yes ○ No

### 7. Miscellaneous / Remarks

7.1 What do you think we can improve? Is there anything else you would like to tell us personally?

(maximum 1000 characters)

THANK YOU VERY MUCH FOR TAKING THE TIME TO PARTICIPATE IN OUR SURVEY.
